# COVID-19 Vaccine Hesitancy in Underserved Communities of North Carolina

**DOI:** 10.1101/2021.02.21.21252163

**Authors:** Irene A. Doherty, William Pilkington, Laurin Brown, Victoria Billings, Undi Hoffler, Lisa Paulin, K. Sean Kimbro, Brittany Baker, Tianduo Zhang, Tracie Locklear, Seronda Robinson, Deepak Kumar

**Author notes:** **Corresponding author:** Deepak Kumar, PhD, Julius L. Chambers Biomedical Biotechnology Research Institute (JLC-BBRI), North Carolina Central University, 700 George Street, Durham, NC 27707.

## Abstract

**Background:** In the United States, underserved communities including Blacks and Latinx are disproportionately affected by COVID-19, and widespread vaccination is critical for curbing this pandemic. This study sought to estimate the prevalence of COVID-19 vaccine hesitancy, describe attitudes related to vaccination, and identify correlates among racial minority and marginalized populations across 9 counties in North Carolina.

**Methods:** We conducted a cross-sectional survey with a self-administered questionnaire distributed at free COVID-19 testing events in underserved rural and urban communities from August 27 – December 15, 2020. Vaccine hesitancy was defined as the response of “no” or “don’t know/not sure” to whether the participant would get the COVID-19 vaccine as soon as it became available.

**Results:** The sample comprised 948 participants including 27.7% Whites, 59.6% Blacks, 12.7% Latinx, and 63% female. Thirty-two percent earned <$20K annually, 60% owned a computer and ∼80% had internet access at home. The prevalence of vaccine hesitancy was 68.9% including 62.7%, 74%, and 59.5% among Whites, Blacks, and Latinx, respectively. Between September and December, the largest decline in vaccine hesitancy occurred among Whites (27.5 percentage points), followed by Latinx (17.6) and the smallest decline was among Black respondents (12.0). 51.2% of the respondents reported vaccine safety concerns, 23.7% wanted others to get of the respondents reported they would trust health care providers with information about the COVID-19 vaccine. Factors associated with hesitancy in multivariable logistic regression included being female (OR=1.90 95%CI[1.36, 2.64]), being Black (OR=1.68 [1.106 2.45]), calendar month (OR=0.76 [0.63, 0.92]), safety concerns (OR=4.28 [3.06, 5.97]), and government distrust (OR=3.57 [2.26, 5.63]).

**Conclusions:** This study reached underserved minority populations in a number of different locations to investigate COVID-19 vaccine hesitancy. We built on existing relationships and further engaged the community, stake holders and health department to provide free COVID-19 testing. This direct approach permitted assessment of vaccine hesitancy (which was much higher than national estimates), distrust, and safety concerns.

**Highlights:**

- This study surveyed 948 adults at COVID-19 testing sites in 9 counties of North Carolina between August 27 and December 15, 2020 where vaccine hesitancy was widespread including 74% in Blacks, 62.7% in Whites and 59.5% in Latinx.
- Vaccine hesitancy declined over time but remained high for Blacks.
- On-site surveys conducted in underserved areas that were paper-based and self-administered permitted reaching adults with no internet (17%), no cell phone (20%), no computer (40%) and yearly incomes less than 20K (31%).
- Widespread vaccine hesitancy in predominately minority communities of NC must be addressed to successfully implement mass COVID-19 vaccination programs.

## Background

The COVID-19 pandemic in the United States has exacerbated deeply rooted socioeconomic and health disparities in historically marginalized populations.^1,2^ Since the coronavirus epidemic started in the US, it has become one of the leading cause of death.^3,4^ COVID-19 incidence is disproportionately higher among both Blacks and Latinx. Racial minorities are more likely to become severely ill, and nearly three times as likely as Whites to die from COVID-19.^5^

In November 2020, Moderna and Pfizer-BioNTech released their findings from randomized trials of COVID-19 vaccines, showing a remarkable 90-95% efficacy.^6,7^ The Food and Drug Administration (FDA) issued Emergency Use Authorizations (EUA) for the Pfizer-BioNTech vaccine on December 10, 2020^8^ and the Moderna vaccine on December 18, 2020.^9^ The Advisory Committee on Immunization Practices (ACIP) updated recommendations for allocating initial supplies with tiered distribution to groups at highest risk first (e.g., health care workers, nursing home residents, and the elderly)^10,11^ before universal distribution. The population effectiveness^12^ to reach herd immunity thresholds for COVID-19 requires an estimated 70% of the population to be vaccinated.^13,14^ To promote vaccine uptake and access requires resources, strategies and structural intervention in multiple sectors.^15^ Vaccine hesitancy is defined as a “delay in acceptance or refusal of vaccination despite availability of vaccination services.”^16^ The decision to accept, delay, or refuse vaccination is complex depending heavily on the context, place, and specific vaccine. It is paramount that we understand COVID-19 vaccine hesitancy in historically marginalized populations (HMPs) where underlying trust issues directly impact vaccine decisions.

Since early in the pandemic, several studies have used commercial panels, address lists, or random telephone surveys, representative of the US population to estimate vaccine hesitancy.^17–29^ The advantage of panel studies is that a large sample can be enrolled within a very short time (weeks or days), but participation often requires internet access. Two studies conducted during May and June, drawing from different panels, each estimated that 31% of the US population had vaccine hesitancy.^23,27^ Others from approximately the same period reported rates as low as 11% to 25%.^20,21,26^ These studies consistently reported higher levels of vaccine hesitancy among female and Black respondents but it decreased with increasing age. Other indicators of low socioeconomic status (e.g., education and income) were also frequently associated with vaccine hesitancy. Serial polls and studies from the Pew Research Center and others show that vaccine hesitancy has declined around the time the FDA issued EUAs for the two COVID-19 vaccines.^30–32^ Although these studies provide important national estimates of vaccine hesitancy for the overall population and within subpopulations, online surveys do not capture the variants associated with social context, particularly for rural communities with low and intermittent internet connectivity.^33^

In response to the COVID-19 public health emergency, North Carolina Central University (NCCU), a historically black college and university (HBCU), established the <u>A</u>dvanced <u>C</u>enter for <u>CO</u>VID-19 <u>R</u>elated <u>D</u>isparities (ACCORD). ACCORD aims to facilitate COVID-19 testing, multidisciplinary research, and messaging directed at historically marginalized populations in nine North Carolina counties. A key component of the ACCORD efforts is working in close collaboration with community partners. Unlike online incentivized panels, the ACCORD study investigated vaccine hesitancy in targeted predominately rural and urban communities with entrenched, persistent, socioeconomic and health inequalities. The ACCORD strategy was to enroll a convenience sample at COVID-19 testing events hosted by NCCU. This targeted engagement with communities permitted directly assessing the prevalence and identifying correlates of vaccine hesitancy and whether it changed over time.

## Methods

North Carolina has 100 counties that vary widely with respect to population density, rurality and urbanicity, race/ethnicity, socioeconomic, and health indicators. As an HBCU, NCCU has fostered trusting and collaborative relationships with Black underserved communities for decades. Building on these partnerships, ACCORD facilitated COVID-19 testing and surveying programs in nine counties that represent economically-distressed and Health Research Services Administration (HRSA)-designated medically-underserved areas. ACCORD identified local residents as community facilitators and leveraged existing health resources such as public health departments in each county to garner community support for COVID-19 testing events. ACCORD has hosted 52 testing events at locations carefully selected by community facilitators to provide access in otherwise COVID-19 testing deserts. Testing events occurred between August 27, 2020 and December 15, 2020.

ACCORD COVID-19 testing events took place in the parking lots of churches, schools, and similar venues that accommodated drive-through testing. ACCORD partnered with health departments and other service providers to collect nasal swabs for PCR tests. Eligibility criteria to participate in the survey included being at least 18 years of age, English or Spanish comprehension and providing informed consent. NCCU students, faculty and staff greeted individuals in their cars and explained that the university was conducting a survey to better understand the experiences and thoughts about COVID-19 in their community. If they agreed, they received the consent form and survey on a sanitized clipboard with a new ink pen that had never been used (to keep). (Initially, participants were offered sanitized tablets to enter their responses electronically but this was discontinued for logistical reasons.) All volunteers had their temperatures recorded upon arrival, wore masks at all times, sanitized their hands frequently, and used sanitized clipboards and tablets. Survey participants received a variety of NCCU-branded items [e.g., T-shirts, string bags, and cups]) and their names were entered into a monthly raffle for gift cards. The Institutional Review Board at NCCU approved the study.

### Measures

To reduce respondent burden, participants received one of three survey questionnaires that each assessed different topics related to COVID-19 in greater depth. The different surveys assessed psychosocial stress, barriers to COVID-19 testing, contact tracing acceptability, and electronic media use. Each version included a set of core questions. Each survey was clearly marked with a version number (i.e., 1, 2, 3) to ensure that they were evenly distributed. All three versions assessed COVID-19 vaccine hesitancy and one version had additional questions about trusted sources for information about the vaccine.

Vaccine hesitancy was assessed (across all three questionnaire versions) with the question “*Scientists are working on a COVID-19 vaccine. Would you get vaccinated against COVID-19 as quickly as possible when the vaccine becomes available?*” Response choices included *yes, no*, and *don’t know/not sure*. To explore features of vaccine hesitancy, regardless of participants’ responses, the survey then asked, “*Which of the reasons below would stop you or delay you from getting vaccinated against COVID-19 as soon as the vaccine becomes available?*” Participants could choose multiple responses including: 1) *Don’t believe that vaccines work;* 2) *Have concerns about vaccine safety;* 3) *Do not trust the government about the vaccine;* 4) *Do not trust the medical system; and* 5) *Want others to get the vaccine first*.

The outcome used for regression analysis - vaccine hesitancy - combined the responses *no*, and *don’t know/not sure*. Participants who skipped or declined to answer the question were excluded. This analysis also assessed temporal trends for vaccine hesitancy over the data collection period by segmenting the events weeks into three time periods: August-September, October, and November-December. The month number during which the event occurred was treated as a continuous variable in regression models.

### Analysis

All analyses were conducted using Stata Ver 15 (College Station, TX). The analysis generated descriptive statistics and tabular analysis with Chi Square or Fisher’s exacts tests, estimated the prevalence of vaccine hesitancy, quantified reasons to delay or not get vaccinated, and changes over time. Respondents who skipped or declined to answer question(s) were excluded from most analyses. Logistic regression models were used to identify correlates associated with vaccine hesitancy. We selected variables for the adjusted regression model on the basis of previous studies, *a prior* hypotheses and sample size. As a sensitivity analysis, we used general linear models (glm) with a poisson distribution, logit link function, and robust standard errors. The effect estimates were similar and do not change interpretation.

## Results

We recruited a convenience sample of 1,004 participants from 34 testing events. Of these, 948 (94%) reported their race/ethnicity and remained in the analysis. The majority of participants were Black (59.1%), followed by Whites (26.6%) and Latinx (14.4%) (Table 1). Females comprised 63.9% of the sample. The median age for Blacks was 57 and significantly older than Whites (45) and Latinx (37). While disparities in socioeconomic status between Whites and Blacks were evident, Latinx participants were markedly poorer. Significantly more Latinx (40%) did not complete high school or earn a GED as compared to 3.6% of Whites and 9.7% of Blacks. Most respondents lived in a house (68.6%), with noteworthy disparities; 83.5%, 68%, and 42.9% of Whites, Blacks and Latinx living in a house, respectively. Furthermore, 37.9% of Latinx lived in a mobile home as compared to 11.6% of Blacks and 9.3% of Whites. The proportion of Blacks who were married or cohabitating was lowest (39.3%). The majority of Latinx (71.1%) lacked health insurance as compared to Whites (16.7%) and Blacks (14.2%). Approximately half (51%) of Latinx, 30.8% of Blacks and 23.2% of Whites reported an annual income less than $20K. Overall, 15.6%, 38.2%, and 13.1% of respondents reported difficulty paying for food, monthly bills and medical care or prescriptions, respectively.

**Table 1.**
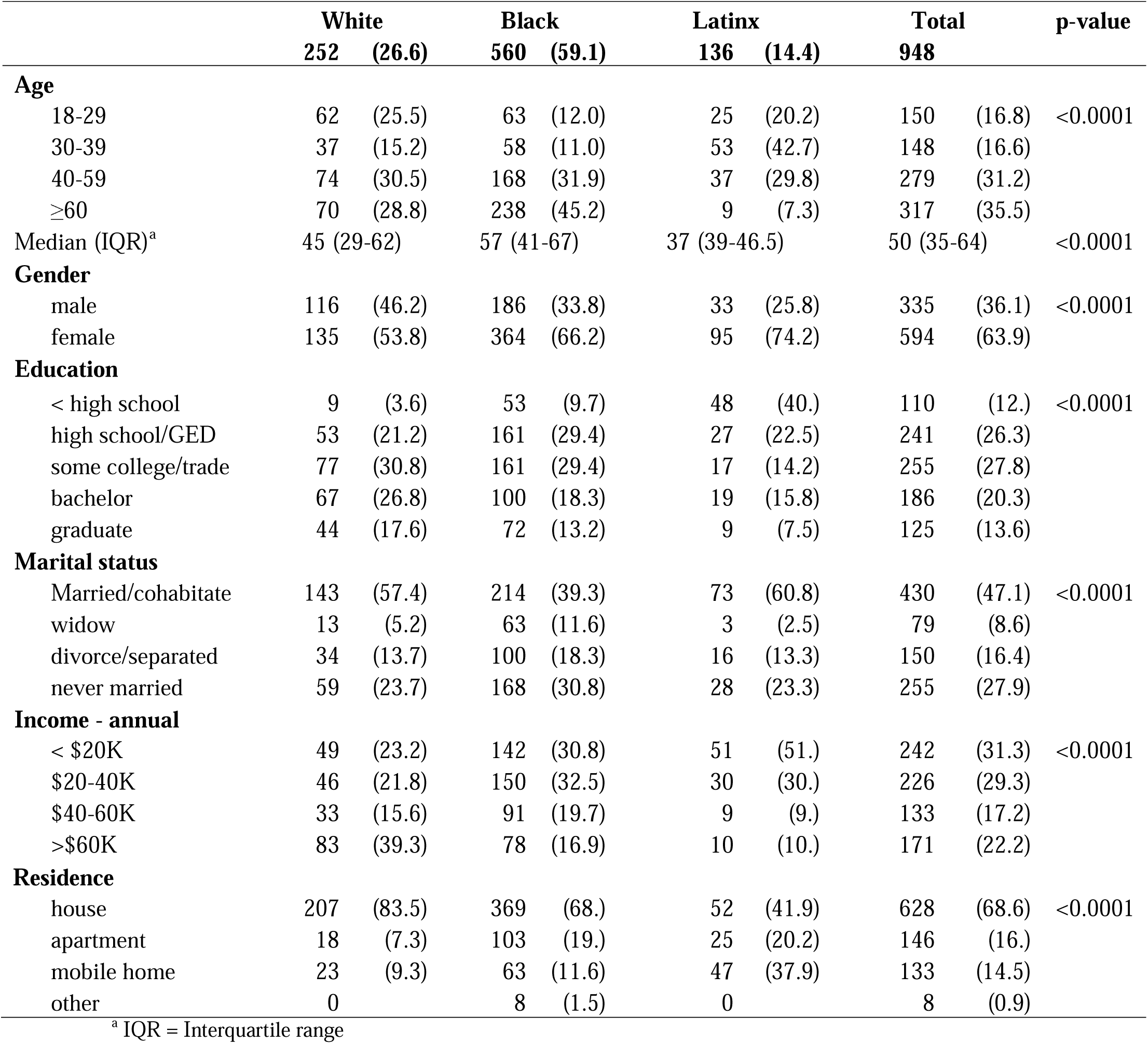
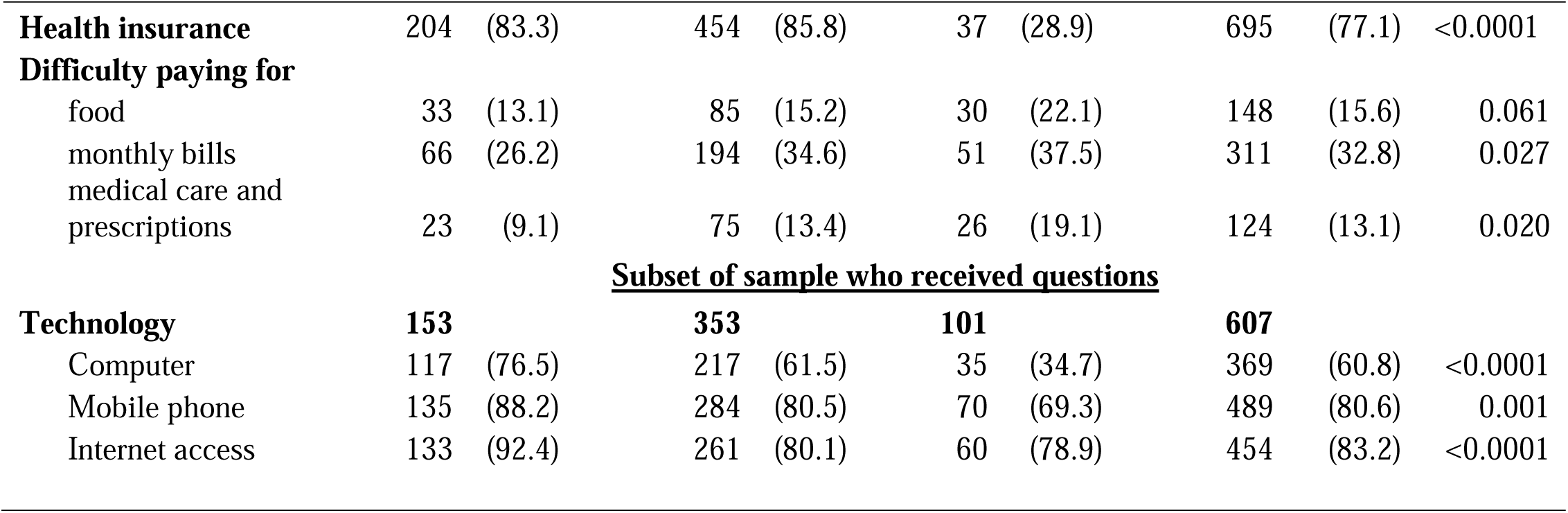
Characteristics of ACCORD study respondents by race/ethnicity ACCORD study, August-December 2020

Disparities with respect to technology and access to the internet was evident among all survey participants. Approximately 39% of respondents did not own a computer. Only 69.3% of Latinx owned a mobile phone as compared 88.2% of Whites and 80.5% of Blacks. Although internet access in the home was nearly universal among Whites (92.4%), significantly fewer Blacks (80.1%) and Latinx (78.7%) reported internet access.

### Vaccine hesitancy

The prevalence of vaccine hesitancy varied by race/ethnicity (Table 2); only 23.4% of Blacks as compared to 36.1% of Whites reported they would get vaccinated as soon as possible. Comparable proportions across all groups (35-36%) were unsure or did not know if they would get it as soon as possible and 89 respondents (9%) skipped the question. Notably, 10% of Blacks and 18.5% of Latinx did not answer the question. Excluding participants who skipped the question and combining the responses *no* and *don’t know* classifies 62.7% of Whites, 74% of Blacks, and 59.5% of Latinx as vaccine hesitant (Table 2). Overall, 69% of respondents reported vaccine hesitancy.

**Table 2.** Prevalence and features of vaccine hesitancy stratified by race, ACCORD Study, North Carolina August – November 2020

| Total n by race | Total sample |  |  |  |  |  |  |  | p-value |
| --- | --- | --- | --- | --- | --- | --- | --- | --- | --- |
|  | White |  | Black |  | Latinx |  | Total |  |  |
|  | 276 | (27.7) | 594 | (59.6) | 126 | (12.7) | 996 | (100.) |  |
|  | N | col% | N | col% | N | col% |  |  |  |
| Get COVID-19 vaccine as soon as possible? |  |  |  |  |  |  |  |  |  |
| yes | 91 | (36.1) | 131 | (23.4) | 44 | (32.6) | 266 | (28.1) | <0.0001 |
| no | 64 | (25.4) | 177 | (31.6) | 17 | (12.6) | 258 | (27.2) |  |
| don't know | 89 | (35.3) | 196 | (35.) | 49 | (36.3) | 334 | (35.3) |  |
| not answered | 8 | (3.2) | 56 | (10.) | 25 | (18.5) | 89 | (9.4) |  |
| Vaccine hesitancy <sup>a,b</sup> | 153 | (62.7) | 373 | (74.0) | 66 | (59.5) | 592 | (68.9) | <0.0001 |
| Reasons for not or delay getting vaccinated |  |  |  |  |  |  |  |  |  |
| doesn't believe vaccines |  |  |  |  |  |  |  |  |  |
| work | 6 | (2.4) | 47 | (8.4) | 6 | (4.4) | 59 | (6.2) |  |
| safety concerns | 137 | (54.4) | 296 | (52.9) | 43 | (31.9) | 476 | (50.3) | <0.0001 |
| mistrust government | 45 | (17.9) | 160 | (28.6) | 18 | (13.3) | 223 | (23.5) | <0.0001 |
| mistrust medical system | 5 | (1.9) | 38 | (14.7) | 7 | (2.1) | 50 | (5.8) |  |
| want others to get vaccine |  |  |  |  |  |  |  |  |  |
| first | 57 | (22.6) | 144 | (25.7) | 23 | (17.) | 224 | (23.7) |  |
| Subset of sample who received questions |  |  |  |  |  |  |  |  |  |
| Total n | 84 | (25.4) | 194 | (58.6) | 53 | (16.) | 331 | (100) |  |
| Trusted sources for vaccine information |  |  |  |  |  |  |  |  |  |
| Health websites | 27 | (36.) | 50 | (26.7) | 8 | (15.4) | 85 | (27.1) | 0.01 |
| Healthcare provider | 53 | (70.7) | 126 | (67.4) | 19 | (36.5) | 198 | (63.1) | <0.0001 |
| Community service |  |  |  |  |  |  |  |  |  |
| organization | 21 | (28.) | 42 | (22.5) | 10 | (19.2) | 73 | (23.2) |  |
| Pastor or other faith leaders | 5 | (6.7) | 18 | (9.6) | 6 | (11.5) | 29 | (9.2) |  |
| Received flu shot within 12 |  |  |  |  |  |  |  |  |  |
| months | 34 | (45.9) | 74 | (42.) | 16 | (39.) | 124 | (42.6) |  |

<sup>a</sup> Excludes n=89 participants who did not answer question
<sup>b</sup> Hesitancy combines responses *no* and *don't know/not sure*

The most common reason for not- or delaying-getting vaccinated was safety concerns for 54,4% of Whites, 52.9% of Blacks, and 31.9% of Latinx (Table 2). Only 17% of Latinx wanted others to receive the vaccine first, as compared to 22.6% of Whites and 25.7% of Blacks. Significantly more Blacks (28.6%) mistrusted the government as compared to 17.9% and 13.3% of Whites and Latinx, respectively. The questionnaire also included a question that referred to mistrust of the medical system that very few participants indicated (5.5%). Among the subset of respondents (n=311) who received additional questions, trusted sources for information about the vaccine diverged significantly for Latinx. For example, only 36.5% of Latinx trust health care providers for information on COVID-19 as opposed to 70.7% and 67.4% of Whites and Blacks, respectively.

Figure 1 presents the distributions of reasons to prevent or delay vaccination stratified by participants’ responses to getting the vaccine as soon as possible. These results reveal important subtleties. Although 28.1% of respondents reported that they would indeed get the vaccine (Table 2), they nevertheless reported they had safety concerns (30.7%) (Figure 1). Respondents who reported they would *not* get vaccinated were also more likely to report that vaccines don’t work (14.7%) and government mistrust (43%). It is important to note that between 24-26% of participants, independent of their vaccine hesitancy (or lack of) wanted others to get the vaccine first (p-value= 0.769).

**Figure 1.**
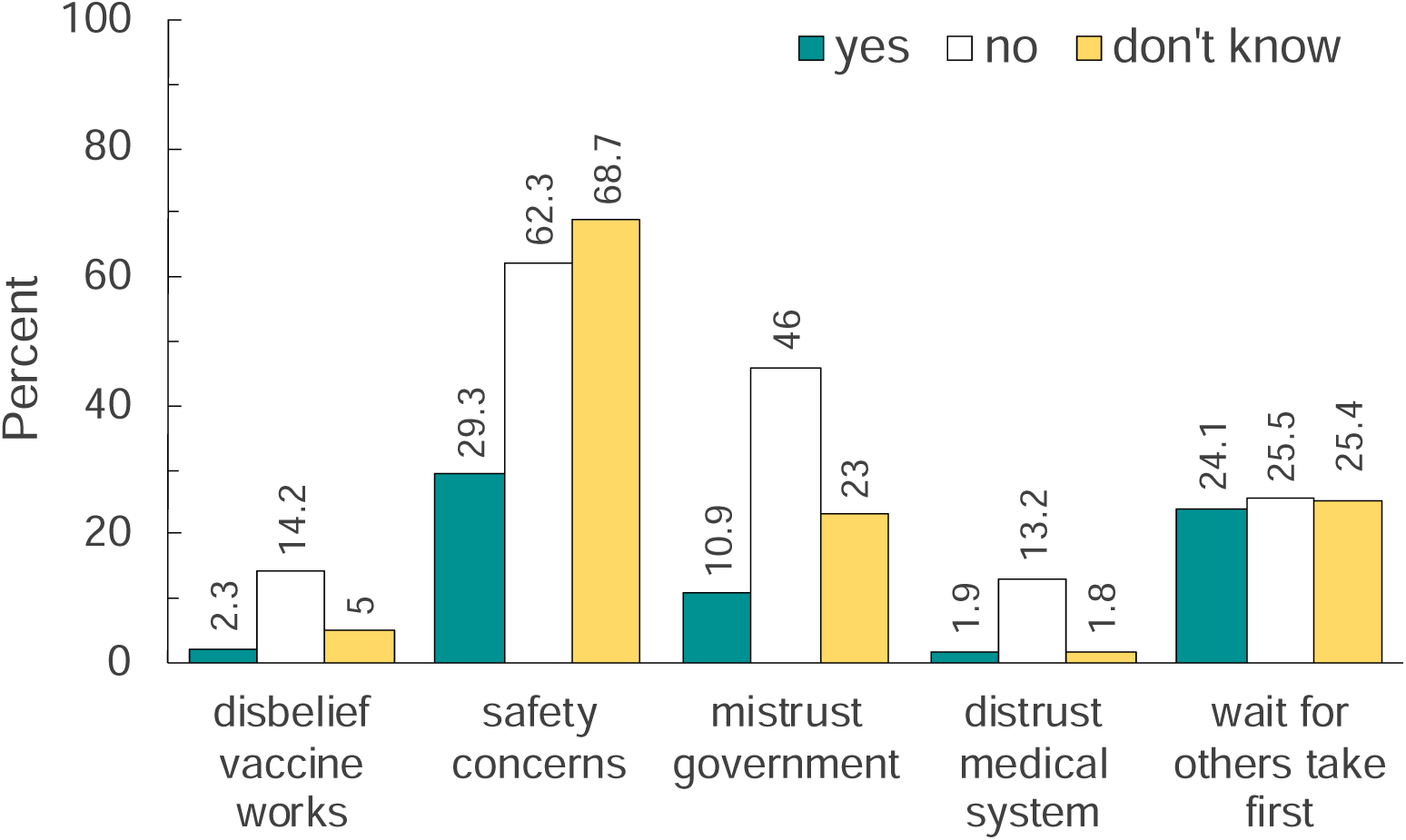
Percentage distribution of reasons to prevent or delay vaccination stratified by whether participants responded that they would get the vaccine as soon as it became available.

Figure 2 displays patterns of temporal changes in three intervals by vaccine hesitancy and stratified by race. The prevalence of hesitancy declined for each group. It was not statistically significant for Latinx because the majority enrolled at testing events during October located at Latino churches in one county. The prevalence of hesitancy among Whites declined by 27.5 percentage points from a high of 69.2% during September to 41.7% during November-December (p-value=0.01). While vaccine hesitancy also significantly declined among Blacks, the prevalence was very high during the first interval (78.4%) and only declined by 12 percentage points (66.4%) during November-December.

**Figure 2.**
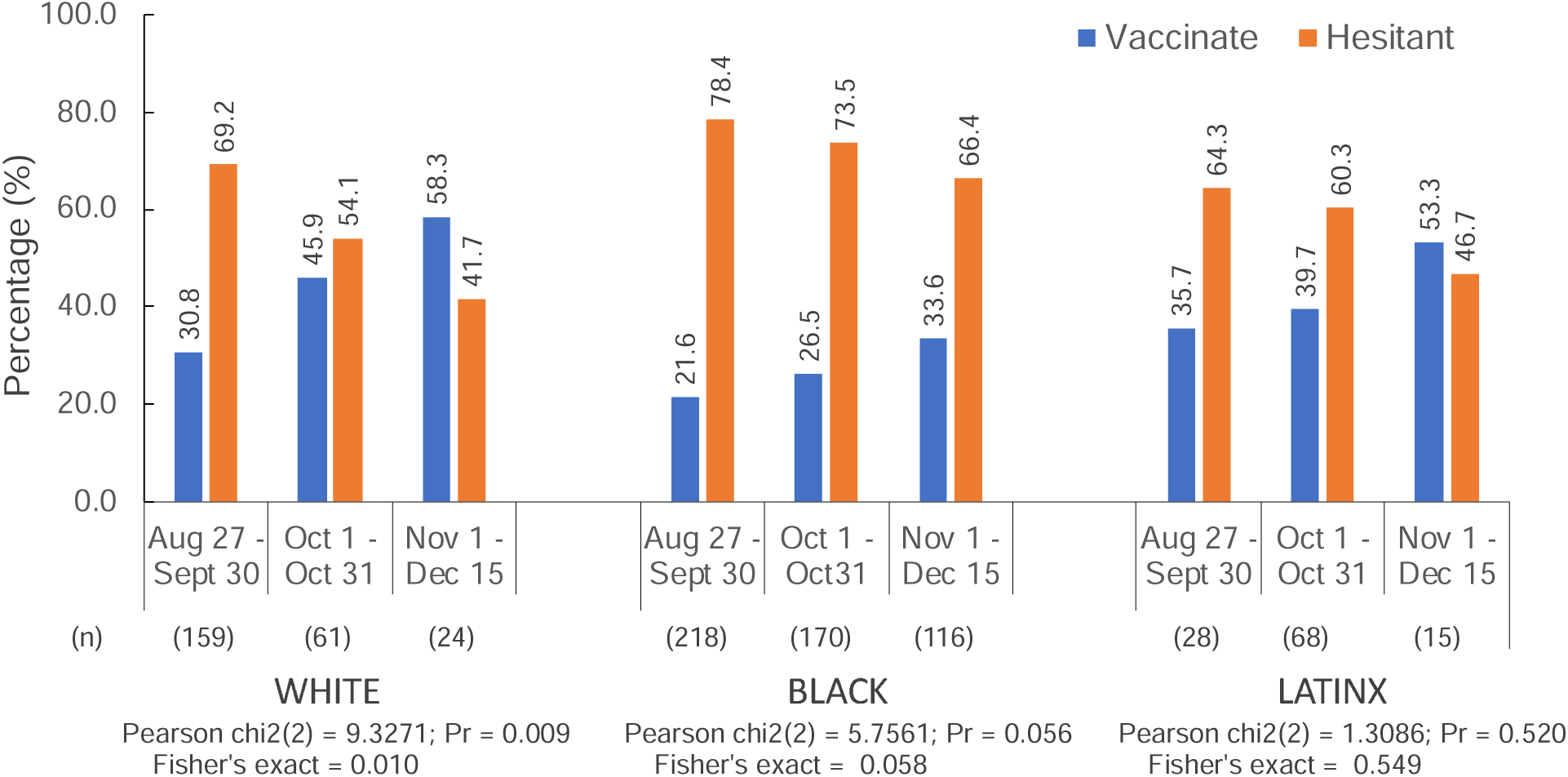
Percentage of US adults (White, Black and Latinx) who will take the COVID-19 vaccine as soon as it becomes available. Those who said yes were classified as Vaccinate and those who said no or were unsure were classified as Hesitant.

### Logistic regression

Table 3 presents the bivariate associations with vaccine hesitancy. Although ownership of mobile phones (OR=2.12, 95%CI[1.31, 3.43]) and computers (OR=1.46, 95%CI[1.00, 2.13]), 3.43] were associated with hesitancy, they were not included in the adjusted model because the questions were asked to a subset of participants (n=607, 64% of sample). In the multivariable model (Table 4), Blacks were 1.68(1.16, 2.45) and females were 1.90 (1.36, 2.64) times as likely to report vaccine hesitancy, as well as participants who expressed safety concerns (OR=4.28 [3.06, 5.97]), government distrust (OR=3.57 [2.26, 5.63]) or wanting others to get vaccinated first (OR=1.44, [0.98, 2.11], marginally significant p=0.062). Hesitancy also persisted to significantly decline over time with each passing month (OR=0.76 [0.63, 0.92]).

**Table 3.** Bivariate Logistic Regression - Vaccine Hesitancy, ACCORD Study, NC August-December 2020

|  | Vaccinate |  | Hesitant |  | Total |  |  |  |  |
| --- | --- | --- | --- | --- | --- | --- | --- | --- | --- |
|  | N | row% | N | row% | N | OR | (95% CI) |  | p-value |
| Race |  |  |  |  |  |  |  |  |  |
| White | 91 | (37.3) | 153 | (62.7) | 244 | ref |  |  |  |
| Black | 131 | (26.0) | 373 | (74.0) | 504 | 1.69 | (1.22 | 2.35) | 0.002 |
| Latinx | 45 | (40.5) | 66 | (59.5) | 111 | 0.87 | (0.55 | 1.38) | 0.560 |
| Gender |  |  |  |  |  |  |  |  |  |
| male | 124 | (39.9) | 187 | (60.1) | 311 | ref |  |  |  |
| female | 141 | (26.2) | 398 | (73.8) | 539 | 1.87 | (1.39 | 2.52) | <0.0001 |
| Data collection period |  |  |  |  |  |  |  |  |  |
| Aug-Sept | 106 | (26.2) | 299 | (73.8) | 405 | ref |  |  |  |
| October | 100 | (33.4) | 199 | (66.6) | 299 | 0.71 | (0.51 | 0.98) | 0.036 |
| Nov-Dec | 61 | (39.4) | 94 | (60.6) | 155 | 0.55 | (0.37 | 0.81) | 0.002 |
| Event month (8-12) (continuous variable) |  |  |  |  |  | 0.79 | (0.67 | 0.93) | 0.004 |
| Age |  |  |  |  |  |  |  |  |  |
| 18-29 | 45 | (31.5) | 98 | (68.5) | 143 | ref |  |  |  |
| 30-39 | 42 | (31.8) | 90 | (68.2) | 132 | 0.98 | (0.59 | 1.64) | 0.950 |
| 40-59 | 60 | (23.7) | 193 | (76.3) | 253 | 1.48 | (0.94 | 2.33) | 0.094 |
| ≥60 | 100 | (35.1) | 185 | (64.9) | 285 | 0.85 | (0.55 | 1.30) | 0.456 |
| per 10 year age increase (continuous variable) |  |  |  |  |  | 0.93 | (0.86 | 1.01) | 0.106 |
| Education |  |  |  |  |  |  |  |  |  |
| < high school | 31 | (35.6) | 56 | (64.4) | 87 | ref |  |  |  |
| high school/GED | 66 | (30.8) | 148 | (69.2) | 214 | 1.24 | (0.73 | 2.10) | 0.420 |
| some college/trade | 75 | (31.1) | 166 | (68.9) | 241 | 1.23 | (0.73 | 2.05) | 0.441 |
| bachelor | 41 | (23.4) | 134 | (76.6) | 175 | 1.81 | (1.03 | 3.17) | 0.038 |
| graduate | 42 | (35.3) | 77 | (64.7) | 119 | 1.01 | (0.57 | 1.81) | 0.960 |
| Income |  |  |  |  |  |  |  |  |  |
| < 20K | 69 | (32.7) | 142 | (67.3) | 211 | ref |  |  |  |
| 20-40K | 61 | (28.6) | 152 | (71.4) | 213 | 1.21 | (0.80 | 1.83) | 0.365 |
| 40-60K | 28 | (21.9) | 100 | (78.1) | 128 | 1.74 | (1.04 | 2.88) | 0.034 |
| >60k | 54 | (32.9) | 110 | (67.1) | 164 | 0.99 | (0.64 | 1.53) | 0.963 |
| Marital Status |  |  |  |  |  |  |  |  |  |
| married/cohabitate | 129 | (33.3) | 258 | (66.7) | 387 | ref |  |  |  |
| widow | 22 | (31.) | 49 | (69.) | 71 | 1.11 | (0.65 | 1.92) | 0.699 |
| divorce/separated | 40 | (29.2) | 97 | (70.8) | 137 | 1.21 | (0.79 | 1.85) | 0.374 |
| never married | 65 | (27.3) | 173 | (72.7) | 238 | 1.33 | (0.93 | 1.90) | 0.115 |
| Residence |  |  |  |  |  |  |  |  |  |
| house | 175 | (30.2) | 405 | (69.8) | 580 | ref |  |  |  |
| apartment | 39 | (29.8) | 92 | (70.2) | 131 | 1.02 | (0.67 | 1.54) | 0.928 |
| mobile home | 44 | (36.7) | 76 | (63.3) | 120 | 0.75 | (0.49 | 1.13) | 0.163 |
| other | 2 | (28.6) | 5 | (71.4) | 7 | 1.08 | (0.21 | 5.62) | 0.927 |

Table 3 continued
|  | Vaccinate |  | Hesitant |  | Total |  |  |  |  |
| --- | --- | --- | --- | --- | --- | --- | --- | --- | --- |
|  | N | row% | N | row% | N | OR | (95% CI) |  | p-value |
| **Health insurance** |  |  |  |  |  |  |  |  |  |
| Yes | 200 | (30.8) | 450 | (69.2) | 650 | ref |  |  |  |
| No | 61 | (33.5) | 121 | (66.5) | 182 | 0.88 | (0.62 | 1.25) | 0.480 |
| **Difficulty paying for:** |  |  |  |  |  |  |  |  |  |
| food | 42 | (30.7) | 95 | (69.3) | 137 | 1.02 | (0.69 | 1.52) | 0.907 |
| monthly bills | 84 | (29.5) | 201 | (70.5) | 285 | 1.12 | (0.82 | 1.53) | 0.473 |
| medical care or prescriptions | 32 | (28.6) | 80 | (71.4) | 112 | 1.15 | (0.74 | 1.78) | 0.538 |
| **Pandemic** |  |  |  |  |  |  |  |  |  |
| Know someone who had COVID | 106 | (28.2) | 270 | (71.8) | 376 | 1.25 | (0.93 | 1.68) | 0.143 |
| **Reasons to prevent or delay vaccine** |  |  |  |  |  |  |  |  |  |
| doesn't believe vaccines work | 7 | (12.1) | 51 | (87.9) | 58 | 3.50 | (1.57 | 7.82) | 0.002 |
| safety concerns | 82 | (17.6) | 383 | (82.4) | 465 | 4.13 | (3.03 | 5.64) | <0.0001 |
| mistrust government | 29 | (13.7) | 183 | (86.3) | 212 | 3.67 | (2.41 | 5.61) | <0.0001 |
| want others to get vaccine first | 64 | (29.1) | 156 | (70.9) | 220 | 1.13 | (0.81 | 1.59) | 0.459 |
| **Technology** |  |  |  |  |  |  |  |  |  |
|  |  |  |  |  |  |  | **<u>Subset of sample who received questions (n=607)</u>** |  |  |
| internet access | 122 | (28.0) | 313 | (72.0) | 435 | 1.28 | (0.78 | 2.10) | 0.322 |
| mobile phone | 126 | (27.0) | 341 | (73.0) | 467 | 2.12 | (1.31 | 3.43) | 0.002 |
| computer | 97 | (26.8) | 265 | (73.2) | 362 | 1.46 | (1.00 | 2.13) | 0.053 |

**Table 4.** Multivariable Logistic Regression – Vaccine Hesitancy ACCORD Study, NC August-December 2020

|  | **OR** | **(95% CI)** | **p-value** |
| --- | --- | --- | --- |
| **Race** |  |  |  |
| White | ref |  |  |
| Black | 1.68 | (1.16 2.45) | 0.006 |
| Latinx | 1.14 | (0.67 1.91) | 0.634 |
| **Gender** |  |  |  |
| male | ref |  |  |
| female | 1.90 | (1.36 2.64) | <0.0001 |
| **Calendar time** |  |  |  |
| Event month (8-12) | 0.76 | (0.63 0.92) | 0.004 |
| **Reasons to prevent or delay vaccine** |  |  |  |
| safety concerns | 4.28 | (3.06 5.97) | <0.0001 |
| mistrust government | 3.57 | (2.26 5.63) | <0.0001 |
| want others to get vaccine first | 1.44 | (0.98 2.11) | 0.062 |

## Discussion

The ACCORD study enrolled over 1000 participants attending COVID-19 testing events that took place in targeted communities with high racial/ethnic minorities which suffer from health disparities and economic inequalities. The pandemic has intensified historically valid distrust of the government, medical establishment, and probably academic researchers. Our success in conducting this study emanates from the innate trust embedded in the definition of an HBCU, in addition to enlisting local residents as community facilitators who looked like participants, and our on-the-ground approach.

One other study in the US (to date) mirrors our methods.^34^ Researchers at the University of California Berkeley have collaborated with Latino farmworker communities for decades. They recruited 1115 farmworkers receiving free COVID-19 tests at clinics and other community events during July – November 2020. Half (52%) of participants reported they would be extremely likely to get vaccinated and 32% were unsure. Most vaccine-hesitant participants reported worry about side effects (∼65%). While this study and the ACCORD study reached different marginalized populations, they share similar concerns. Both studies suggest that working within communities where trust is earned will be essential for addressing vaccination and the many needs that the COVID-19 pandemic has created or made worse.

Unlike representative online panel studies,^17–28,35,36^ we captured vast differences in COVID-19 vaccine hesitancy among historically marginalized populations living in areas that lack technology and reliable internet service.^33^ The estimated prevalence of vaccine hesitancy in national studies ranged between ∼11% to 35% whereas in the ACCORD study, it ranged between 42% to 79% depending on the race/ethnicity and collection window. Some online studies collected political party affiliation or conservative versus liberal attitudes to quantify the partisan impacts on pandemic response.^19,22,28,37,38^ In general, White, Republicans, and conservatives were less likely to wear masks or practice social distancing. Fortunately, Dr. Anthony Fauci has become a household name and research suggests that his endorsement of wearing masks and COVID-19 vaccines increases confidence and uptake among Democrats and Republicans alike.^28^

Nevertheless, these findings reflect the charged social climate during the four months of the ACCORD study. This period, August-December 2020, will remain prominently ingrained in US history for reasons other than the pandemic. Shelter-in-place orders caused widespread economic hardships, Black Lives Matter and the death of George Floyd instilled more activism, the incidence of COVID-19 fluctuated across the country, a new president was elected, and two highly efficacious COVID-19 vaccines^6,7^ received emergency use authorizations from the FDA. Meanwhile, attitudes about the pandemic and vaccines, in particular, are rapidly changing. Repeated polls report that vaccine hesitancy has dropped over time (yet remains higher among Blacks).^31,39^ Surveys of health care workers reported more hesitancy (than the public) at the beginning of the pandemic, but as the results from clinical trials were released, hesitancy diminished precipitously.^40,41^

Despite these promising trends, vaccine hesitancy may continue to persist among historically marginalized populations unless substantial resources, funding, messaging, public health activities, and access to vaccination programs are deployed. Intervention research scientists often engage African American pastors and church leaders to promote healthy behaviors.^42,43^ Our findings show that fewer than 10% of respondents seek the advice of the church for COVID-19 vaccine information. Still, the importance and influence of faith leaders, however, cannot be undermined. ignored or diminished. Although, the majority of respondents (62.5%) placed their trust in their health care providers, efforts must be made to collectively utilize community and faith leaders and health providers of color^24^ to deliver accurate and reliable information about the vaccine.^15^ Misinformation may change attitudes and the appearance of any side effects from the first shot may deter people from receiving their second shot and any booster vaccinations. Providing transparent information about vaccine development, potential side effects, and answering related questions will help ameliorate vaccine hesitancy.

There are limitations of the ACCORD survey. The data originate from a convenience self-selected sample recruited from COVID-19 testing events who agreed to take the extra time to complete the survey. The extent to which they reflect the experiences of others in their communities is unclear. Because respondents completed paper surveys, we did not have the advantages of electronic data entry including logic checks, skip patterns, and reducing missing data. However, online surveys tended to skip the questions about reasons not to vaccinate. Such questions were administered to all respondents in our paper survey, showing that people who were not vaccinate hesitant, nonetheless, had safety concerns and wanted others to get the vaccine first (Figure 2). Logistical constraints prevented collecting COVID-19 test results from participants. That said, *not* collecting testing results likely increased participation and preserved community trust.

The ACCORD communities were testing deserts and susceptible to becoming vaccination deserts perpetuating health disparities if provision of vaccines fails to address and overcome obstacles. Emerging evidence suggests that navigating the landscape to obtain vaccines requires a combination of information about where to go, health literacy, computer savvy, patience, and persistence.^44,45^ ACCORD presents an example of a coordinated system, where collected meaningful data for the implementing of evolving COVID-19 management and vaccination strategies on-the-ground in most vulnerable and underserved communities.

## Data Availability

Data is available upon request following Data Use Agreement.

## Acknowledgement

We acknowledge community/health partners and NCCU students, faculty and staff who volunteered at the many COVID testing sites organized by NCCU ACCORD.

## Author contributions

Drs. Kumar and Doherty had full access to the data in the study and take responsibility for the integrity of the data, collection and the accuracy of the data analysis.

**Concept and Design:** Kumar, Pilkington and Doherty

**Data acquisition, analysis, or interpretation of data:** All authors

**Drafting of the manuscript:** Doherty, Kumar

**Critical review of the manuscript for important intellectual content:** All authors

**Statistical analysis:** Doherty

**Obtained funding:** Kumar

**Administrative, technical, or material support:** Kumar

**Supervision:** Kumar

## References

1. Webb Hooper M, Nápoles AM, Pérez-Stable EJ. COVID-19 and Racial/Ethnic Disparities. JAMA. 2020;323(24):2466–2467. doi:10.1001/jama.2020.8598

2. Clouston SAP, Natale G, Link BG. Socioeconomic inequalities in the spread of coronavirus-19 in the United States: A examination of the emergence of social inequalities. Soc Sci Med. 2021;268:113554. doi:10.1016/j.socscimed.2020.113554

3. Woolf SH, Chapman DA, Lee JH. COVID-19 as the Leading Cause of Death in the United States. JAMA. Published online December 17, 2020. doi:10.1001/jama.2020.24865

4. Woolf SH, Chapman DA, Sabo RT, Weinberger DM, Hill L, Taylor DDH. Excess Deaths From COVID-19 and Other Causes, March-July 2020. JAMA. 2020;324(15):1562. doi:10.1001/jama.2020.19545

5. CDC. Coronavirus Disease 2019 (COVID-19). Centers for Disease Control and Prevention. Published February 11, 2020. Accessed December 23, 2020. https://www.cdc.gov/coronavirus/2019-ncov/covid-data/investigations-discovery/hospitalization-death-by-race-ethnicity.html

6. Baden LR, El Sahly HM, Essink B, et al. Efficacy and Safety of the mRNA-1273 SARS-CoV-2 Vaccine. N Engl J Med. 2020;0(0):null. doi:10.1056/NEJMoa2035389

7. Polack FP, Thomas SJ, Kitchin N, et al. Safety and Efficacy of the BNT162b2 mRNA Covid-19 Vaccine. N Engl J Med. 2020;383(27):2603–2615. doi:10.1056/NEJMoa2034577

8. Coronavirus (COVID-19) Update: FDA Holds Advisory Committee Meeting to Discuss Authorization of COVID-19 Vaccine Candidate as Part of Agency’s Review of Safety and Effectiveness Data. FDA. Published December 10, 2020. Accessed December 22, 2020. https://www.fda.gov/news-events/press-announcements/coronavirus-covid-19-update-fda-holds-advisory-committee-meeting-discuss-authorization-covid-19

9. FDA Takes Additional Action in Fight Against COVID-19 By Issuing Emergency Use Authorization for Second COVID-19 Vaccine. FDA. Published December 21, 2020. Accessed December 22, 2020. https://www.fda.gov/news-events/press-announcements/fda-takes-additional-action-fight-against-covid-19-issuing-emergency-use-authorization-second-covid

10. Committee on Equitable Allocation of Vaccine for the Novel Coronavirus, Board on Health Sciences Policy, Board on Population Health and Public Health Practice, Health and Medicine Division, National Academies of Sciences, Engineering, and Medicine. Framework for Equitable Allocation of COVID-19 Vaccine. (Gayle H, Foege W, Brown L, Kahn B, eds.). National Academies Press; 2020:25917. doi:10.17226/25917

11. Dooling K. The Advisory Committee on Immunization Practices’ Updated Interim Recommendation for Allocation of COVID-19 Vaccine — United States, December 2020. MMWR Morb Mortal Wkly Rep. 2020;69. doi:10.15585/mmwr.mm695152e2

12. Weinberg GA, Szilagyi PG. Vaccine Epidemiology: Efficacy, Effectiveness, and the Translational Research Roadmap. J Infect Dis. 2010;201(11):1607–1610. doi:10.1086/652404

13. Kwok KO, Lai F, Wei WI, Wong SYS, Tang JWT. Herd immunity – estimating the level required to halt the COVID-19 epidemics in affected countries. J Infect. 2020;80(6):e32–e33. doi:10.1016/j.jinf.2020.03.027

14. Fine P, Eames K, Heymann DL. “Herd Immunity”: A Rough Guide. Clin Infect Dis. 2011;52(7):911–916. doi:10.1093/cid/cir007

15. Wood S, Schulman K. Beyond Politics — Promoting Covid-19 Vaccination in the United States. Malina D, ed. N Engl J Med. Published online January 6, 2021:NEJMms2033790. doi:10.1056/NEJMms2033790

16. MacDonald NE. Vaccine hesitancy: Definition, scope and determinants. Vaccine. 2015;33(34):4161–4164. doi:10.1016/j.vaccine.2015.04.036

17. Pogue K, Jensen JL, Stancil CK, et al. Influences on Attitudes Regarding Potential COVID-19 Vaccination in the United States. Vaccines. 2020;8(4):582. doi:10.3390/vaccines8040582

18. Fisher KA, Bloomstone SJ, Walder J, Crawford S, Fouayzi H, Mazor KM. Attitudes Toward a Potential SARS-CoV-2 Vaccine: A Survey of U.S. Adults. Ann Intern Med. 2020;173(12):964–973. doi:10.7326/M20-3569

19. Kreps S, Prasad S, Brownstein JS, et al. Factors Associated With US Adults’ Likelihood of Accepting COVID-19 Vaccination. JAMA Netw Open. 2020;3(10):e2025594. doi:10.1001/jamanetworkopen.2020.25594

20. Taylor S, Landry CA, Paluszek MM, Groenewoud R, Rachor GS, Asmundson GJG. A Proactive Approach for Managing COVID-19: The Importance of Understanding the Motivational Roots of Vaccination Hesitancy for SARS-CoV2. Front Psychol. 2020;11:575950. doi:10.3389/fpsyg.2020.575950

21. Southwell BG, Kelly BJ, Bann CM, Squiers LB, Ray SE, McCormack LA. Mental Models of Infectious Diseases and Public Understanding of COVID-19 Prevention. Health Commun. 2020;35(14):1707–1710. doi:10.1080/10410236.2020.1837462

22. Carpiano RM. Demographic Differences in US Adult Intentions to Receive a Potential Coronavirus Vaccine and Implications for Ongoing Study. medRxiv. Published online September 9, 2020:2020.09.07.20190058. doi:10.1101/2020.09.07.20190058

23. Reiter PL, Pennell ML, Katz ML. Acceptability of a COVID-19 vaccine among adults in the United States: How many people would get vaccinated? Vaccine. 2020;38(42):6500–6507. doi:10.1016/j.vaccine.2020.08.043

24. Head KJ, Kasting ML, Sturm LA, Hartsock JA, Zimet GD. A National Survey Assessing SARS-CoV-2 Vaccination Intentions: Implications for Future Public Health Communication Efforts. Sci Commun. 2020;42(5):698–723. doi:10.1177/1075547020960463

25. Muñana C, 2020. KFF COVID-19 Vaccine Monitor: December 2020 -Methodology. KFF. Published December 15, 2020. Accessed December 27, 2020. https://www.kff.org/report-section/kff-covid-19-vaccine-monitor-december-2020-methodology/

26. Khubchandani J, Sharma S, Price JH, Wiblishauser MJ, Sharma M, Webb FJ. COVID-19 Vaccination Hesitancy in the United States: A Rapid National Assessment. J Community Health. Published online January 3, 2021. doi:10.1007/s10900-020-00958-x

27. Callaghan T, Moghtaderi A, Lueck JA, et al. Correlates and disparities of intention to vaccinate against COVID-19. Soc Sci Med. Published online January 4, 2021:113638. doi:10.1016/j.socscimed.2020.113638

28. Bokemper SE, Huber GA, Gerber AS, James EK, Omer SB. Timing of COVID-19 vaccine approval and endorsement by public figures. Vaccine. 2021;39(5):825–829. doi:10.1016/j.vaccine.2020.12.048

29. Szilagyi PG, Thomas K, Shah MD, et al. National Trends in the US Public’s Likelihood of Getting a COVID-19 Vaccine—April 1 to December 8, 2020. JAMA. 2021;325(4):396. doi:10.1001/jama.2020.26419

30. Daly M, Robinson E. Willingness to Vaccinate against COVID-19 in the US: Longitudinal Evidence from a Nationally Representative Sample of Adults from April–October 2020. Public and Global Health; 2020. doi:10.1101/2020.11.27.20239970

31. Intent to Get a COVID-19 Vaccine Rises to 60% as Confidence in Research and Development Process Increases. Pew Research Center; 2020. Accessed December 22, 2020. https://www.pewresearch.org/science/2020/12/03/intent-to-get-a-covid-19-vaccine-rises-to-60-as-confidence-in-research-and-development-process-increases/

32. Elon University Poll. Elon University. Accessed January 22, 2021. https://www.elon.edu/u/elon-poll/

33. Digital gap between rural and nonrural America persists Pew Research Center. Accessed December 22, 2020. https://www.pewresearch.org/fact-tank/2019/05/31/digital-gap-between-rural-and-nonrural-america-persists/

34. Mora AM, Lewnard JA, Kogut K, et al. Impact of the COVID-19 Pandemic and Vaccine Hesitancy among Farmworkers from Monterey County, California. medRxiv. Published online January 1, 2020:2020.12.18.20248518. doi:10.1101/2020.12.18.20248518

35. Szilagyi PG, Thomas K, Shah MD, et al. National Trends in the US Public’s Likelihood of Getting a COVID-19 Vaccine—April 1 to December 8, 2020. JAMA. Published online December 29, 2020. doi:10.1001/jama.2020.26419

36. Ruiz JB, Bell RA. Predictors of intention to vaccinate against COVID-19: Results of a nationwide survey. Vaccine. Published online January 2021:S0264410X21000141. doi:10.1016/j.vaccine.2021.01.010

37. Ruiz JB, Bell RA. Predictors of intention to vaccinate against COVID-19: Results of a nationwide survey. Vaccine. 2021;39(7):1080–1086. doi:10.1016/j.vaccine.2021.01.010

38. Khubchandani J, Sharma S, Price JH, Wiblishauser MJ, Sharma M, Webb FJ. COVID-19 Vaccination Hesitancy in the United States: A Rapid National Assessment. J Community Health. Published online January 3, 2021. doi:10.1007/s10900-020-00958-x

39. Muñana C, 2020. KFF COVID-19 Vaccine Monitor: December 2020.; 2020. Accessed December 27, 2020. https://www.kff.org/coronavirus-covid-19/report/kff-covid-19-vaccine-monitor-december-2020/

40. Meyer MN, Gjorgjieva T, Rosica D. Healthcare worker intentions to receive a COVID-19 vaccine and reasons for hesitancy: A survey of 16,158 health system employees on the eve of vaccine distribution. medRxiv. Published online January 1, 2020:2020.12.19.20248555. doi:10.1101/2020.12.19.20248555

41. Shekhar R, Sheikh AB, Upadhyay S, et al. COVID-19 Vaccine Acceptance among Health Care Workers in the United States. Vaccines. 2021;9(2):119. doi:10.3390/vaccines9020119

42. Corbie-Smith G, Goldmon M, Roman Isler M, et al. Partnerships in Health Disparities Research and the Roles of Pastors of Black Churches: Potential Conflict, Synergy, and Expectations. J Natl Med Assoc. 2010;102(9):823–831. doi:10.1016/S0027-9684(15)30680-5

43. Williams RM, Glanz K, Kegler MC, Davis E. A Study of Rural Church Health Promotion Environments: Leaders’ and Members’ Perspectives. J Relig Health. 2012;51(1):148–160. doi:10.1007/s10943-009-9306-2

44. Ianzito C. Frustration Over COVID Vaccine Distribution. AARP. Published January 27, 2021. Accessed January 29, 2021. <http://www.aarp.org/health/conditions-treatments/info-2021/vaccine-distribution.html>

45. Hamil, Liz, Kirziner, Ashley, Lopes L, Sparks, Grace, Brodie, Mollyan. KFF COVID-19 Vaccine Monitor: January 2021.; 2021. Accessed January 29, 2021. https://www.kff.org/coronavirus-covid-19/report/kff-covid-19-vaccine-monitor-january-2021/

